# Risk factors of coronary microvascular obstruction

**DOI:** 10.1101/2020.05.29.20116665

**Authors:** Yong Li, Shuzheng Lyu

## Abstract

**Background:** Coronary microvascular obstruction /no-reflow(CMVO/NR) is a predictor of long-term mortality in survivors of ST elevation myocardial infarction (STEMI) underwent primary percutaneous coronary intervention (PPCI).

**Objective:** To identify risk factors of CMVO/NR during PPCI.

**Methods:** Totally 2384 STEMI patients treated with PPCI were divided into two groups according to thrombolysis in myocardial infarction(TIMI) flow grade:CMVO/NR group(246cases, TIMI 0-2 grade) and control group(2138 cases, TIMI 3 grade). We used univariable and multivariable logistic regression to identify risk factors of CMVO/NR during PPCI.

**Results:** A frequency of CMVO/NR was 10.3%(246/2384). Logistic regression analysis showed that the differences between the two groups in age(unadjusted odds ratios [OR] 1.032; 95% CI, 1.02 to 1.045; adjusted OR 1.032; 95% CI, 1.02 to 1.046; P <0.001), periprocedural bradycardia (unadjusted OR 2.357; 95% CI, 1.752 to 3.171; adjusted OR1.818; 95% CI, 1.338 to 2.471; P <0.001),using thrombus aspiration devices during operation (unadjusted OR 2.489; 95% CI, 1.815 to 3.414; adjusted OR1.835; 95% CI, 1.291 to 2.606; P =0.001),neutrophil percentage (unadjusted OR 1.028; 95% CI, 1.014 to 1.042; adjusted OR1.022; 95% CI, 1.008 to 1.036; P =0.002), and total occlusion of the culprit vessel (unadjusted OR 2.626; 95% CI, 1.85 to 3.728; adjusted-OR 1.656;95% CI, 1.119 to 2.45; P =0.012) were statistically significant (P <0. 05). The area under the receiver operating characteristic curve was 0.6896

**Conclusions:** Age, periprocedural bradycardia, using thrombus aspiration devices during operation, neutrophil percentage, and total occlusion of the culprit vessel may be independent risk factors for predicting CMVO/NR during PPCI.

We registered this study with WHO International Clinical Trials Registry Platform (ICTRP) (registration number: ChiCTR1900023213; registered date: 16 May 2019).http://www.chictr.org.cn/edit.aspx?pid=39057&htm=4.

## Background

Coronary microvascular obstruction /no-reflow(CMVO/NR) is a predictor of long-term mortality in survivors of ST elevation myocardial infarction (STEMI)underwent primary percutaneous coronary intervention (PPCI). ^[1-4]^ CMVO/NR is defined as inadequate myocardial perfusion after successful mechanical opening of the infarct-related artery. ^[1,4,5]^ CMVO/NR is diagnosed immediately after PCI when postprocedural angiographic thrombolysis in myocardial infarction(TIMI) flow is < 3, or in the case of a TIMI flow of 3 when myocardial blush grade is 0 or 1, or when ST resolution within 60–90 min of the procedure is < 70%.^[4,5]^ There have been few large clinical trials of therapies, specifically aimed at reducing CMVO/NR. ^[1,6]^ Prevention of CMVO/NR is a crucial step. We want to identify risk factors of CMVO/NR during PPCI.

## Methods

Totally 2384 STEMI patients who were consecutively treated with PPCI in Beijing Anzhen Hospital, Capital Medical University between 2007 and 2018.

Prior to emergency angiography, all patients received 300 mg of aspirin, 300 to 600 mg of clopidogrel or 180 mg of ticagrelor and unfractionated heparin.

Inclusion criteria: STEMI patients presenting within 12 hours from the symptom onset who were treated with PPCI. We established the diagnosis of acute myocardial infarction (AMI) and STEMI base on fourth universal definition of myocardial infarction. ^[7]^

Exclusion criteria. 1. patients received thrombolysis; 2. patients received bivalirudin.

CMVO/NR was defined as TIMI <3 after successful mechanical opening of the infarct-related arterie with a guide wire, a balloon, or a stent, not the end of the PPCI process. ^[4] [5]^

We selected 15 predictors based on clinical relevance and the results of the pre-experiment cohort. They were shown in Table 1. Periprocedural bradycardia was defined as preoperative heart rate ≥ 50 times / min, intraoperative heart rate <50 times / min persistent or transient. ^[8]^ Intra-procedural hypotension was defined as pre-procedural systolic blood pressure was > 90mmHg, intra-procedural systolic blood pressure less than or equal to 90 mmHg persistent or transient. ^[9]^ Whether a thrombus aspiration device was used depends on interventionalist.

**Table 1.**
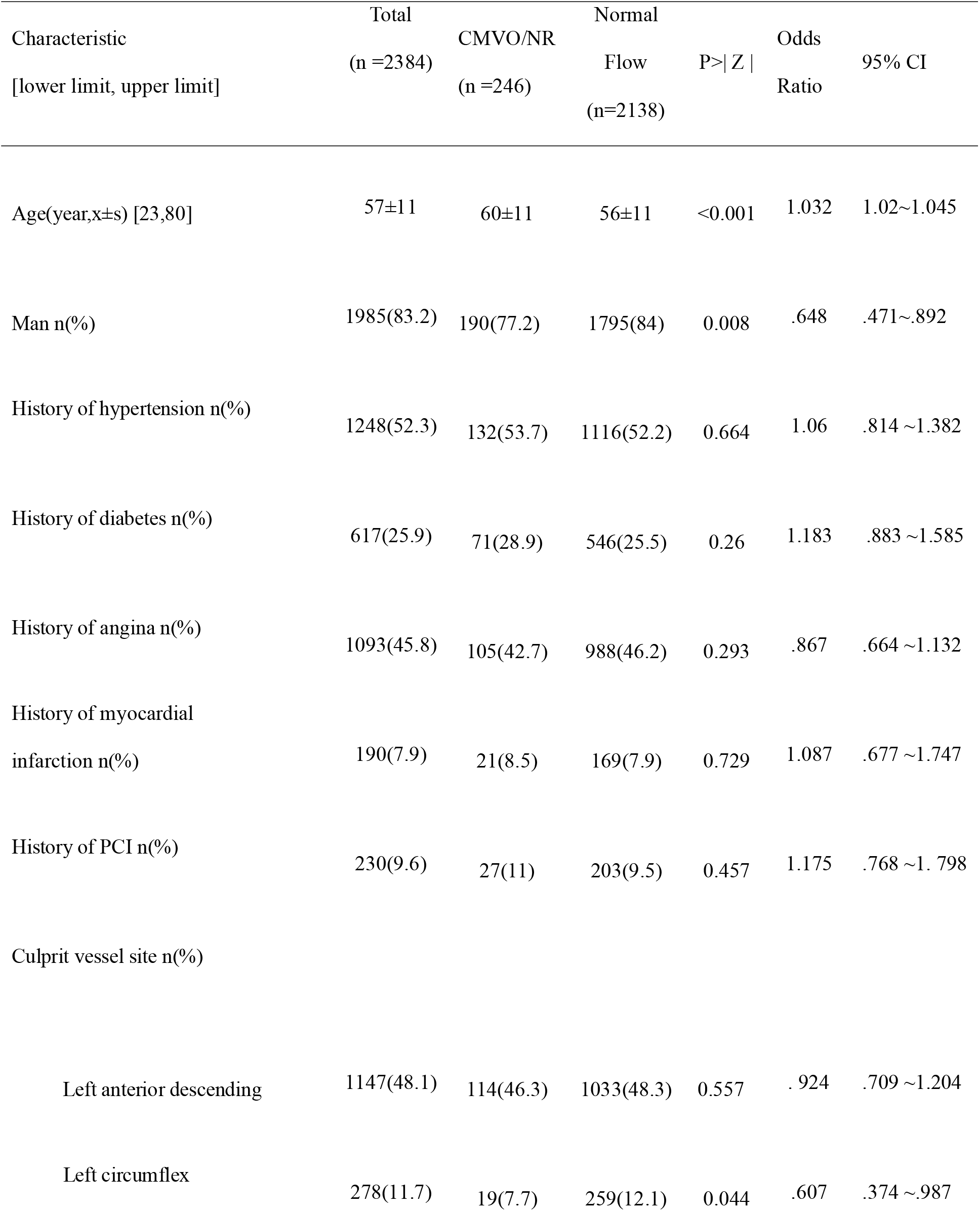
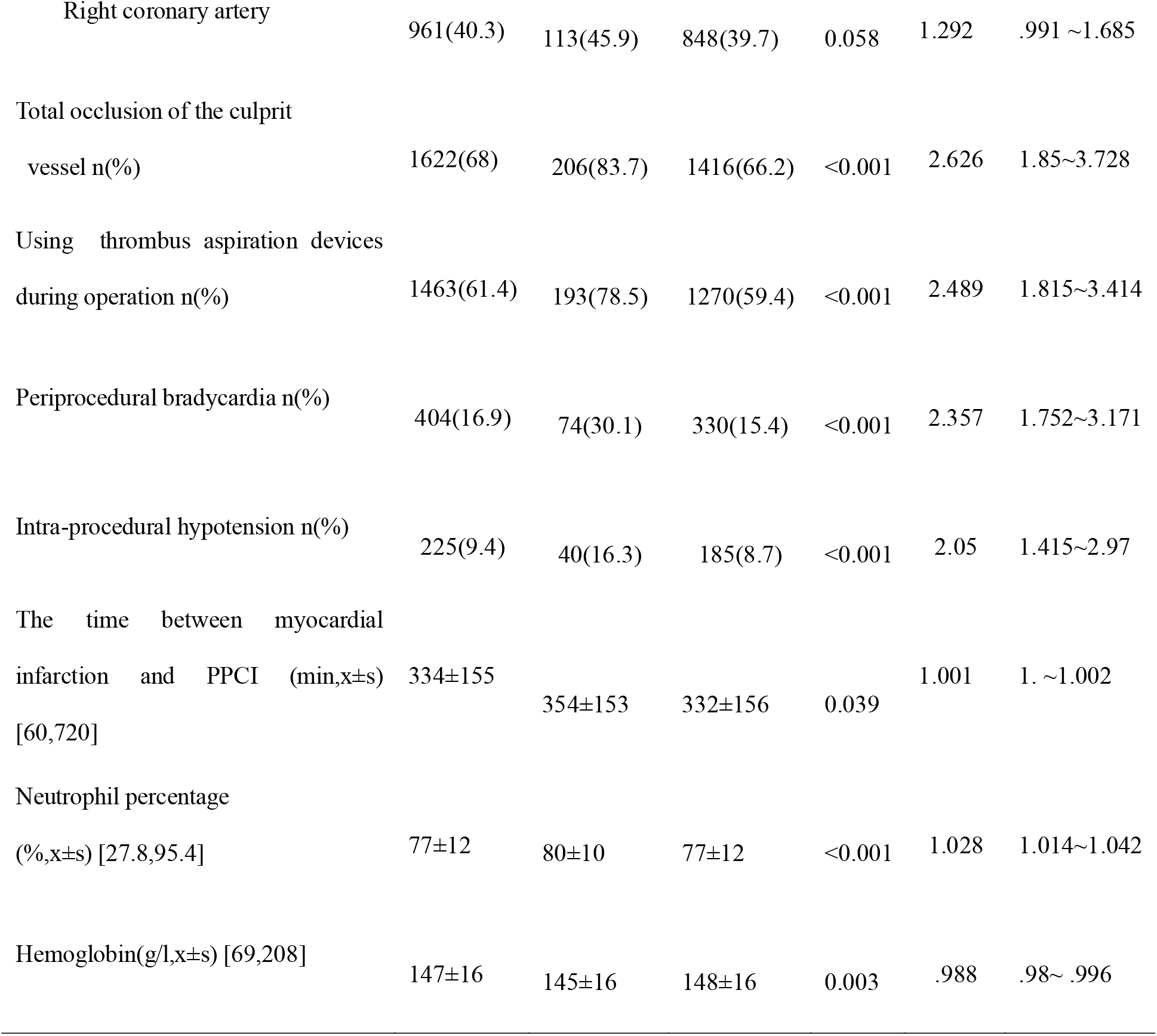
Demographic, clinical, and angiographic characteristics of patients with CMVO/NR and normal coronary flow during PPCI.

| Characteristic<br>[lower limit, upper limit] | Total<br>(n =2384) | CMVO/NR<br>(n =246) | Normal<br>Flow<br>(n=2138) | P>\| Z \| | Odds<br>Ratio | 95% CI |
| --- | --- | --- | --- | --- | --- | --- |
| Age(year,x±s) [23,80] | 57±11 | 60±11 | 56±11 | <0.001 | 1.032 | 1.02~1.045 |
| Man n(%) | 1985(83.2) | 190(77.2) | 1795(84) | 0.008 | .648 | .471~.892 |
| History of hypertension n(%) | 1248(52.3) | 132(53.7) | 1116(52.2) | 0.664 | 1.06 | .814 ~1.382 |
| History of diabetes n(%) | 617(25.9) | 71(28.9) | 546(25.5) | 0.26 | 1.183 | .883 ~1.585 |
| History of angina n(%) | 1093(45.8) | 105(42.7) | 988(46.2) | 0.293 | .867 | .664 ~1.132 |
| History of myocardial<br>infarction n(%) | 190(7.9) | 21(8.5) | 169(7.9) | 0.729 | 1.087 | .677 ~1.747 |
| History of PCI n(%) | 230(9.6) | 27(11) | 203(9.5) | 0.457 | 1.175 | .768 ~1. 798 |
| Culprit vessel site n(%) |  |  |  |  |  |  |
| Left anterior descending | 1147(48.1) | 114(46.3) | 1033(48.3) | 0.557 | . 924 | .709 ~1.204 |
| Left circumflex | 278(11.7) | 19(7.7) | 259(12.1) | 0.044 | .607 | .374 ~.987 |
| Right coronary artery | 961(40.3) | 113(45.9) | 848(39.7) | 0.058 | 1.292 | .991 ~1.685 |
| Total occlusion of the culprit vessel n(%) | 1622(68) | 206(83.7) | 1416(66.2) | <0.001 | 2.626 | 1.85~3.728 |
| Using thrombus aspiration devices during operation n(%) | 1463(61.4) | 193(78.5) | 1270(59.4) | <0.001 | 2.489 | 1.815~3.414 |
| Periprocedural bradycardia n(%) | 404(16.9) | 74(30.1) | 330(15.4) | <0.001 | 2.357 | 1.752~3.171 |
| Intra-procedural hypotension n(%) | 225(9.4) | 40(16.3) | 185(8.7) | <0.001 | 2.05 | 1.415~2.97 |
| The time between myocardial infarction and PPCI (min,x±s) [60,720] | 334±155 | 354±153 | 332±156 | 0.039 | 1.001 | 1. ~1.002 |
| Neutrophil percentage (% ,x±s) [27.8,95.4] | 77±12 | 80±10 | 77±12 | <0.001 | 1.028 | 1.014~1.042 |
| Hemoglobin(g/l,x±s) [69,208] | 147±16 | 145±16 | 148±16 | 0.003 | .988 | .98~ .996 |

### Statistical analysis

We followed the methods of Li et al. 2019. ^[10]^ We constructed a multivariable logistic regression model using the backward variable selection method.Statistical analyses were performed with STATA version 15.1 .P value <0.05 was considered statistically significant.

## Results

During PPCI procedure, 2138 patients had a TIMI flow grade 3 (group with normal epicardial flow) and 246patients had a TIMI flow grade 0~2 (group with CMVO/NR). Baseline characteristics of the patients were shown in Table 1. We used univariable and multivariable logistic regression to identify predictors of CMVO/NR. We identified 10 variables (age, sex, total occlusion of the culprit vessel, periprocedural bradycardia, intra-procedural hypotension, using thrombus aspiration devices during operation, neutrophil percentage, hemoglobin, the time between myocardial infarction and PPCI, and the culprit vessel was left circumflex) as predictors of CMVO/NR in univariable analysis. Five variables (age, periprocedural bradycardia, using thrombus aspiration devices during operation, neutrophil percentage, and total occlusion of the culprit vessel) remained as independent predictors of CMVO/NR in multivariable analysis. The results were shown in Table 2 and Table3. The receiver operating characteristic curve was drawn.The area under the receiver operating characteristic curve was 0.6896±0.017, 95% CI=0.656~0.723.

**Table 2.** Predictor of CMVO/NR obtained from multivariable logistic regression models (odds ratio)

| CMVO/NR | Odds ratio | Std.Err | Z | P>\| Z \| | 95% CI |
| --- | --- | --- | --- | --- | --- |
| Age | 1.032 | .007 | 4.96 | <0.001 | 1.02~1.045 |
| Periprocedural bradycardia | 1.818 | .285 | 3.82 | <0.001 | 1.338~2.471 |
| Using thrombus aspiration devices during operation | 1.835 | .329 | 3.39 | 0.001 | 1.291~2.606 |
| Total occlusion of the<br>culprit vessel | 1.656 | .331 | 2.52 | 0.012 | 1.119~2.45 |
| Neutrophil percentage | 1.022 | 0.007 | 3.14 | 0.002 | 1.008~1.036 |
| Cons | .001 | .001 | -9.64 | <0.001 | .0003~.005 |

**Table 3.** Predictor of CMVO/NR obtained from multivariable logistic regression models (Coef)

| CMVO/NR | Coef | Std.Err | Z | P>\| Z \| | 95% CI |
| --- | --- | --- | --- | --- | --- |
| Age | .032 | .006 | 4.96 | <0.001 | .019~.045 |
| Periprocedural<br>bradycardia | .598 | .156 | 3.82 | <0.001 | .291~.905 |
| Using thrombus<br>aspiration devices<br>during operation | .607 | .179 | 3.39 | 0.001 | .256~.958 |
| Total occlusion of the<br>culprit vessel | .504 | .2 | 2.52 | 0.012 | .112 ~.896 |
| Neutrophil percentage | .022 | .007 | 3.14 | 0.002 | .008 ~.035 |
| Cons | -6.677 | .692 | -9.64 | <0.001 | -8.034~-5.32 |

## Discussion

CMVO/NR is a multifactorial phenomenon and five mechanisms have been recognized: (i) pre-existing microvascular dysfunction, (ii) distal microthrombo-embolization, (iii) ischemic injury, (iv) reperfusion injury and (v) individual susceptibility. ^[5]^ In our study, age, periprocedural bradycardia, using thrombus aspiration devices during operation, neutrophil percentage, and total occlusion of the culprit vessel are associated with an increased risk of CMVO/NR.

Advanced age has been reported to be an independent risk factor of CMVO/NR. ^[5]^ Previous studies had shown that abnormal non-endothelium-dependent microvasodilation appears to be related to functional and structural changes that lead to impaired coronary blood flow reserve with aging.^[5]^

Periprocedural bradycardia may be a sign of CMVO/NR. ^[10]^ In our study, patients with periprocedural bradycardia were at 1.82 higher risk of CMVO/NR than patients without periprocedural bradycardia. Myocardial reperfusion can evoke activation of Bezold-Jarisch reflex.^[11]^ The Bezold-Jarisch reflex means bradycardia, vasodilation, and hypotension. ^[12]^ Acetylcholine, which is endothelium-dependent vasodilator, induces coronary dilation in young healthy subjects but cause vasoconstriction in patients with atherosclerosis. ^[10]^ Excessive vagus nerve excitation is an important factor that may cause CMVO/NR. We should inhibit it to prevent and treat CMVO/NR.

Total occlusion of the culprit vessel is a independent risk factor of CMVO/NR. ^[13]^ In our study, patients with total occlusion of culprit vessel were at 1.66 higher risk of CMVO/NR than patients without total occlusion of the culprit vessel. Good patency of the infarct-related artery prior to PPCI suggests lower thrombus burden and so on. ^[14]^

Neutrophil percentage is an independent risk factor for CMVO/NR. Neutrophil plugging plays a role in the pathogenesis of CMVO/NR. A massive infiltration of microcirculation by neutrophils occurs at the time of reperfusion. ^[15]^ Activated neutrophils release reactive oxygen species and proinflammatory molecules, which can contribute to CMVO/NR. ^[15]^

Using thrombus aspiration device during PCI is closely related to CMVO/NR. Routine thrombus aspiration is not recommended, but in cases of large residual thrombus burden after opening the vessel with a guide wire or a balloon, thrombus aspiration may be considered. ^[4]^ When the thrombus load is high, the thrombus aspiration device tends to be used while CMVO/NR tends to occur.

### Study Limitations

This is a single center experience. Some patients were enrolled >10 years ago thus their treatment may not conform to current standards and techniques. Only TIMI flow grade was used to identify CMVO/NR, and no other diagnostic methods were used because of limited data. We want to get risk factors of CMVO/NR before it happen, some variables associated with CMVO/NR was not including, so the area under the receiver operating characteristic curve was not strong enough.

## Conclusions

Age, periprocedural bradycardia, using thrombus aspiration devices during operation, neutrophil percentage, and total occlusion of the culprit vessel may be independent risk factors for predicting CMVO/NR during PPCI.

List of abbreviations
AMI: Acute myocardial infarction
CMVO: Coronary microvascular obstruction
NR: No-reflow
PCI: Percutaneous coronary intervention
PPCI: Primary percutaneous coronary intervention
STEMI: ST elevation myocardial infarction
TIMI: Trombolysis in myocardial infarction risk score

## Declarations

### Ethics approval and consent to participate

Ethic committee approved the study. Approved No. of ethic committee: 2019013X.

Name of the ethic committee : Ethics committee of Beijing Anzhen Hospital Capital Medical University.It was a retrospective analysis and informed consent was waived by Ethics Committee of Beijing Anzhen Hospital Capital Medical University.

### Statement of human and animal rights

All procedures performed in studies involving human participants were in accordance with the ethical standards of the institutional and/or national research committee and with the 1964 Helsinki declaration and its later amendments or comparable ethical standards. The study was not conducted with animals.

### Consent for publication

Not applicable.

### Availability of data and materials

The data used to support the findings of this study are included within the supplementary information file.

The data are demographic, clinical, and angiographic characteristics of patients with acute STEMI during PPCI. AH= History of angina; AGE= age; TOCV=total occlusion of the culprit vessel; CNR=CMVO/NR; DH= history of diabetes; H=hemoglobin; HH = history of hypertension; IH= introperative hypotension; LAD= the culprit vessel was left anterior descending coronary artery; LCX= the culprit vessel was left circumflex coronary artery;MIH=history of myocardial infarction; NP=neutrophil percentage; PB=periprocedural bradycardia; PCIH=history of percutaneous coronary intervention; RCA= the culprit vessel was right coronary artery; S = sex; TA=using thrombus aspirationdevices during operation; TBMIPPCI =the time between myocardial infarction and PPCI.

## Data Availability

The data used to support the findings of this study are included within the supplementary information file.
The data are demographic, clinical, and angiographic characteristics of patients with acute STEMI during PPCI. AH= History of angina; AGE= age; TOCV=total occlusion of the culprit vessel; CNR=CMVO/NR; DH= history of diabetes; H=hemoglobin; HH = history of hypertension; IH= introperative hypotension; LAD= the culprit vessel was left anterior descending coronary artery; LCX= the culprit vessel was left circumflex coronary artery; MIH=history of myocardial infarction; NP=neutrophil percentage; PB=periprocedural bradycardia; PCIH=history of percutaneous coronary intervention; RCA= the culprit vessel was right coronary artery; S = sex; TA=using thrombus aspirationdevices during operation; TBMIPPCI =the time between myocardial infarction and PPCI.

https://pan.baidu.com/s/1mD8NvdLqWr7zQ270o8VzDg

## Authors’ contributions

Yong Li contributed to generating, analysing, and interpreting the study data and drafted the manuscript. Shuzheng Lyu contributed to planning and revised the manuscript critically for important intellectual content. Yong Li and Shuzheng Lyu are responsible for the overall content as guarantor. All authors have read and approved the manuscript.

## Acknowledgements

None.

